# Improved assessment of *Schistosoma* community infection through data resampling methodology

**DOI:** 10.1101/2023.09.26.23296164

**Authors:** David Gurarie, Anirban Mondal, Martial L Ndeffo-Mbah

## Abstract

**Introduction:** The conventional diagnostic for *Schistosoma mansoni* infection is stool microscopy with Kato-Katz technique to detect eggs. Its outcomes are highly variable on day-to-day basis, and may lead to biased estimates of community infection used to inform public health programs. Our goal is to develop a resampling methodology that leverages data from a large-scale randomized trial to accurately predict community infection.

**Methods:** We developed a resampling methodology that provides unbiased community estimates of prevalence, intensity and other statistics for *S. mansoni* infection when a community survey is conducted using single Kato-Katz stool microscopy per host. It leverages a large-scale dataset, collected in the SCORE project, and allows linking single-stool community screening to its putative multi-day ‘true statistics’.

**Results:** SCORE data analysis reveals limited sensitivity of Kato-Katz stool microscopy, and systematic bias of single-day community testing vs. multi-day testing; for prevalence estimate, it can fall up to 50% below true value. The proposed SCORE-cluster methodology reduces systematic bias and brings estimated prevalence values within 5-10% of the true value. This holds for a broad swath of transmission settings, including SCORE communities, and other datasets.

**Discussion:** Our SCORE-cluster methodology can markedly improve the *S. mansoni* prevalence estimate in settings using stool microscopy.

## Introduction

Schistosomiasis is one of the most prevalent neglected tropical diseases with over 250 million people affected worldwide [1], and the collective burden estimated at 3.3 million DALYs (disability-adjusted life years lost) [2]. Intensified control efforts have over the past 15 years focused on preventive chemotherapy via mass drug administration (MDA) with Praziquantel [2-3]. Accurate assessment of community infection, alongside (demographic) risk factors and spatial-temporal environmental patterns, are expected to provide essential inputs to guide control interventions.

Commonly used tools for community assessment employ Kato-Katz (KK) egg-count diagnostics for *S. mansoni*, and urine filtration for *S. haemotobium* [7]. Both are notoriously uncertain with high day-to-day variability of egg-counts for individual hosts [1-4]. Furthermore, a sizable fraction of repeated tests, combine ‘zero’ and ‘positive’ counts for the same individual. So, a single test could qualify a host as ‘positive’ or ‘negative’, which results in highly uncertain and variable outcomes on the community level. Having a multi-stool host screening, like SCORE dataset (Schistosomiasis Consortium for Operational Research and Evaluation), can achieve far higher accuracy [6], but such tests could be prohibitively expensive. Realistic control-surveillance programs often rely on single-slide/host screening. Such screening can greatly underestimate community infection.

There were several proposals to address diagnostic variability and uncertainty of KK tests via statistical modeling, [3, 5] [6, 7], and mathematical models [8-10]. These models make certain assumptions about putative worm burden distribution in a host community, the ensuing egg release and KK screening, all expressed through parametric distributions, like negative binomial et al. Fitting model to data one can infer unknown parameters and outputs (prevalence, intensity) from model analysis.

Here we propose an alternative empirical approach that utilizes resampling of multi-slide SCORE community tests and statistical inferences drawn from data analysis. Each community resample consists of random draws from mixed tests. Such resample can be viewed as a putative community snapshot, to be observed in a typical control/surveillance setting. Our goal is to apply the SCORE dataset to arbitrary single-slide community test data (SCORE or non-SCORE) and estimate its unknown true prevalence-intensity statistics along with their uncertainties (error-margins). The analysis of resampled data revealed that single-day KK diagnostics consistently underestimates the multi-day ‘true’ prevalence, and had a wide range of variability (error margins). We observed this pattern across a broad swath of SCORE communities’.

The proposed methodology and computing tools allow to recover true community statistics and reduce prediction uncertainty, by identifying a suitable cluster of SCORE community tests for a given sample test.

To validate our methodology, we applied it to a wide range of single-slide resamples drawn from SCORE and non-SCORE datasets for which multi-day testing ‘truth’ available. It showed our scheme could produce robust, statistically reliable predictions with reduced error margins.

## Methods

### A. SCORE dataset

The SCORE project is a large-scale control-surveillance study conducted in several endemic S. *mansoni* areas over the 5-year period [11-14]. The dataset contains 450 endemic communities from Kenya, Cote d’Ivoire and Tanzania. The communities were split into different control arms with annual screening, 3-5 tests/community depending on its arm. These community tests (1744) constitute the data core for our analysis.

The dataset contained 233102 individual tests (single, double, or triple), by the number of stool samples collected from hosts on different days (up to 3). Each stool sample had two 42-mg thick smears, whose egg-counts reported as (A1, B1) – day 1, (A2,B2) etc. We combined two slides (A,B) into a single daily score A+B. The SCORE KK-values differ from the standard EPG (egg per gram) by factor 12.

The bulk of tested subjects were school-age children considered a sentinel pool for community assessment; adults were added in some tests, but their contribution was marginal, so we dropped them. The aggregate SCORE numbers are given in Table 1.

**Table 1:** Individual test statistics.

|  | SINGLE | DOUBLE | TRIPLE | TOTAL |
| --- | --- | --- | --- | --- |
| CHILD | 52824 | 16066 | 141138 | 210028 |
| ADULT | 22655 | 140 | 279 | 23074 |
| ALL | 75479 | 16206 | 141417 | 233102 |

One application of aggregate data includes estimates of KK test sensitivity (Supplement A, Table S1 & S2). Our primary goal here is the community level assessment. The crucial feature of SCORE exploited in our modeling is large number of mixed tests (double, triple) administered to individual hosts. Each random draw from a mixed test can be viewed as a community snapshot – putative outcome of a typical single-slide screening.

### B. Prevalence and Geometric Mean intensity for community assessment

We want to infer prevalence-intensity statistics from a collection of mixed (multi-stool) tests. The conventional approach consists of replacing each multiple count, by its arithmetic mean,

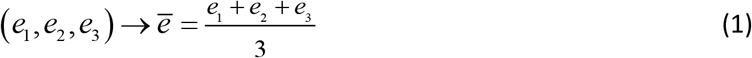

(see e.g. [7, 15], [14, 16, 17]). The resulting community statistics, based on mean scores will be considered ‘true’ values.

Our modeling methodology makes extensive use of two community statistics: conventional prevalence P (fraction of positive counts), and infection intensity measured by the *geometric mean* G of positive counts. The choice of G can be justified on mathematical and empirical grounds (see Supplement B, Figure S2). Two statistics, P-G are largely independent in a broad range of P-values, as demonstrated by SCORE data analysis below

Our goal is to explore variability of single-slide community tests, and their P-G-statistics, in relation to their putative (multi-slide) truth. We do it via extensive analysis of the SCORE dataset. A SCORE community-test would typically combine singlets, doublets, and triplets (1, 2, or 3-day testing), whose ‘true’ values could have different statistical significance. One way to account for significance factor (singlet vs. doublet vs. triplet) is to assign them different weights. Another approach, adopted here, is to confine analysis to triplets alone (dropping the rest). Indeed, triplets comprise the bulk of SCORE tests, about 2/3 (see Table 1). Furthermore, they dominate test data among well-sampled communities (pool size 50 ≤ *m* ≤ 120), as illustrated Figure 1. The resulting collection of community tests (SCORE testbed) are not meant to reproduce realistic SCORE communities, but to serve a basis for the SCORE-cluster methodology developed below.

**Figure 1:**
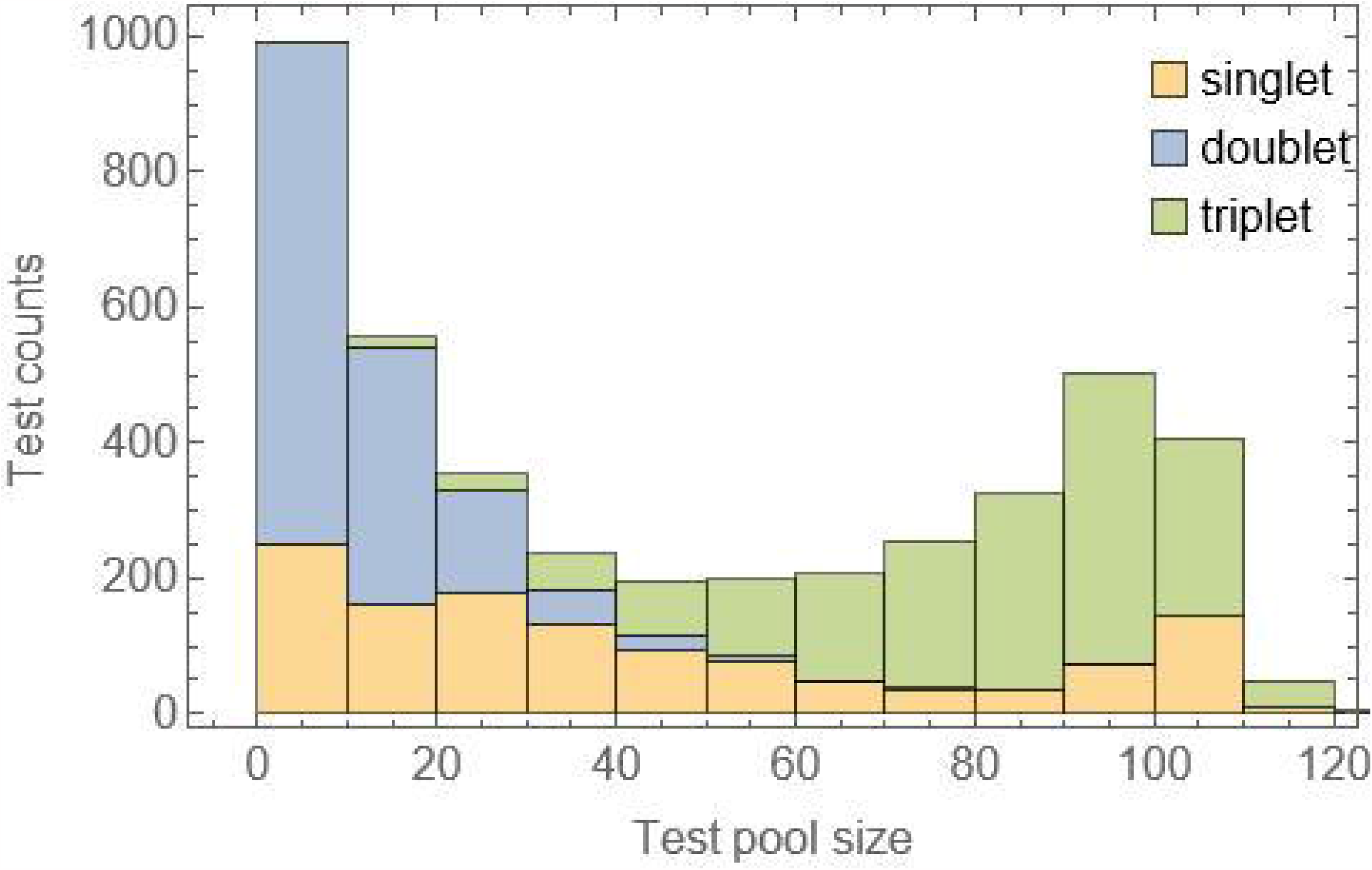
Test-multiplicity distribution by test-pool size. Triplets dominate well-sampled communities (pool size >50)

### C. Resampling methodology for SCORE testbed

The SCORE testbed is confined to young ages (1-15) and triplet test samples. Altogether, we get 1624 community tests, representing a broad range of infection levels, from near zero (*P* ≈ 0.01) to fully infected (*P* ≈ 0.99).

We first examine variability of single-slide KK-diagnostics by generating random resamples (snapshots) of testbed communities. Then we develop a prediction methodology by linking such resamples to ‘truth’.

Each community resample (a single-slide random snapshot) gives a point *z*_*i*_ = (*P*_*i*_, *G*_*i*_) in the prevalance-intensity (PG) plane. An ensemble of resamples makes up a cloud, *C* = {*z*_*i*_ : *i* = 1, 2,…}, with center *z*_0_ = (*P*_0_,*G*_0_) -expected PG-values. We approximate such cloud by a normal distribution *D*_0_ (*z*) = *N* (*z*_0_, Σ_0_) with mean *z*_0_ and covariance matrix Σ_0_. These resample clouds and their distrubutions play crucial role in our methodology.

### D. SCORE-cluster methodology

The main goal is for a given single-slide community snapshot *T* ={*e*_1_, *e*_2_,..} to reconstruct its putative multi-slide SCORE ‘truth’. Raw data T could represent a resampled SCORE community, or a test data collected elsewhere. In either case, we aim to identify a cluster of the SCORE testbed {*T*_*j*_} that are ‘similar’ to *T*, in terms of test statistics. We start by computing PG-values of *T*, reference point *z*_0_ = (*P*_0_, *G*_0_), and for a SCORE cluster {*T*_*j*_} in the testbed, that are likely to reproducing *z*_0_ via resampling.

The proposed procedure, called *SCORE-cluster selection* identifies such pool. It employs a collection of normal distributions {*D*_*j*_ (*z*)} made of 1624 triple SCOREs {*T*_*i*_} -the testbed. Each *D*_*i*_ (*z*) is evaluated at the reference point *z*_0_, and a few high likelihood choices {*w*_*j*_ = *D*_*j*_ (*z*_0_) : *j* = 1,…, *m*} (highest w_j_)are selected as local SCORE-cluster of *z*_0_. Those are most liekely to generate snapshot *T* (in terms of PG-statistics) via random resalpling. Likelihood values{*w*_*j*_} serve as weights of a virtual ‘SCORE-like composite community’ consistent with *T*. We shall use such weighted SCORE-clusters {*T*_*i*_, *w*_*i*_} to infer unknwon true statistics of any raw data *T*. In particular, we estimate its ‘true PG’ values, *Z*_*0*_ =(*P*_*T*_ *G*_*T*_ ), by taking weighted mean ‘cluster truth’ {*Z*_*i*_} .

## Results

### E. KK uncertainty and sensitivity

Mixed SCORE tests exhibit wide day-to-day variability of egg-counts for individual hosts (supplement **Error! Reference source not found**.). Furthermore, a significant portion of mixed tests combines ‘zero’ and ‘positive’ counts (panel (c)). So, the standard binary classification (positive-negative) based on a single KK has reduced sensitivity (Supplement Table S1). Overall, our bulk estimates of KK-sensitivity are consistent with known results (e.g. [7, 15]), but they have little practical use at community level.

### F. Community level analysis

Next, we applied raw-resample methodology across the SCORE triple testbed. Figure 2 (a) illustrates a distribution of 1624 cloud centers, shown as linked mesh-region in the (P, G)-plane. Each resample ensemble was generated from 500 random snapshots from a triplet community test *T*. Cloud centers cover a broad swath in PG-plane. It is reasonable to assume they represent all putative ‘single-slide’ snapshots (SCORE or non-SCORE). Panel (b) shows a typical normal cloud distribution *D*_0_ (*z*) around its marked center *z*_0_ = (*P*_0_,*G*_0_) = (0.41, 2.22). An arrow drawn from *z*_0_ to the SCORE truth *z*_*T*_ = (*P*_*T*_, *G*_*T*_), illustrates the discrepency between snapshot and the

**Figure 2:**
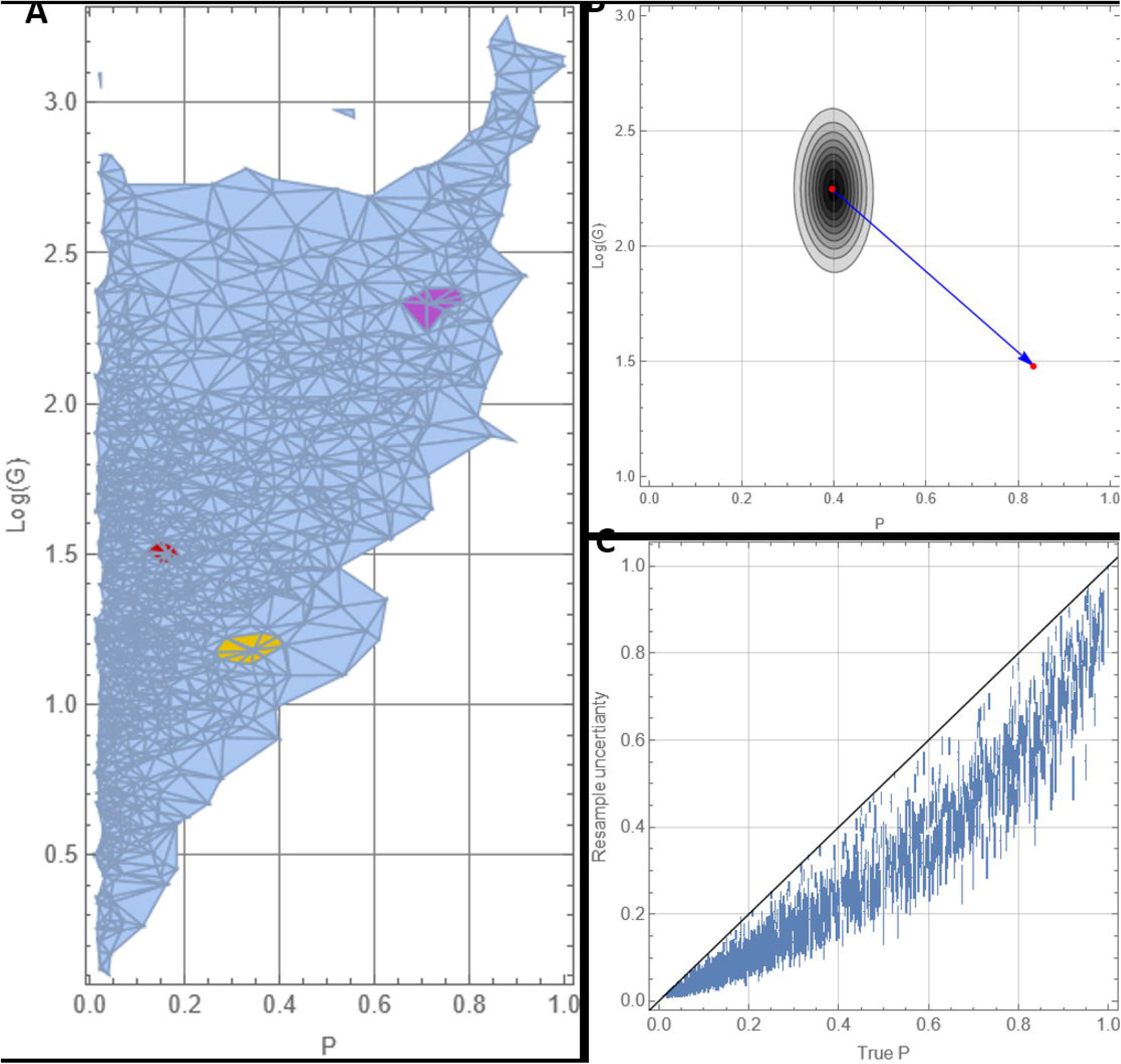
(a) Resample cloud centers of 1624 SCORE community tests in PG-plane, as nodes of the mesh-region with 3 highlighted local SCORE clusters; (b) a PG-cloud approximated by normal distribution *D*_0_ (*z*) centered at *z*_0_ = (0.41, 2.22), with arrow linking *z*_0_ to its multi-slide SCORE ‘truth’ *z*_*T*_ = (0.85,1.48). Panel (c) shows prevalence distributions (bars = mean + SD) of 1624 community tests vs. their SCORE true values (*P*_*T*_).

‘true’ statistics. In this case, true *P*_*T*_ ≈ 0.85 exceeds expected resample *P*_0_ ≈ 0.41, by factor 2.

Panel (c) shows distribution of resampled P-values, mean ± SD (vertical bars) plotted against true *P*_T_ (horizontal axis) across 1624 community tests. We observe a consistent downward bias of resampled P-values, relative to ‘truth’, and wide dispersal within vertical (error) bars.

Our goal is to bridge the gap between ‘truth’ and ‘observation’ by the SCORE-cluster methodology.

### G. Local SCORE-clusters and synthetic composite communities

We employ a testbed of 1624 SCORE clouds, centered at *z* _*j*_ = (*P*_*j*_, *G*_*j*_), and their normal distributions{*D*_*j*_ (*z*)}. Given a single-slide raw-data *T*, and its reference PG-values *z*_0_ = (*P*_0_, *G*_0_), we select a local SCORE cluster of *z*_0_, based on likelihood weights {*w*_*j*_ = *D*_*j*_ (*z*_0_) : *j* = 1,…, *m*}, as explained above. In most applications, 10 highest choices are used (*m* = 10). Figure 2(a) shows highlighted local clusters on the ‘SCORE-center mesh’ for 3 selected reference points.

Figure 3 (a) illustrates local SCORE-cluster of 6 for a red reference point *z*_0_ with 3 ‘highest likelihood’ cloud distributions *D*_j_ along with their w-weights.

**Figure 3:**
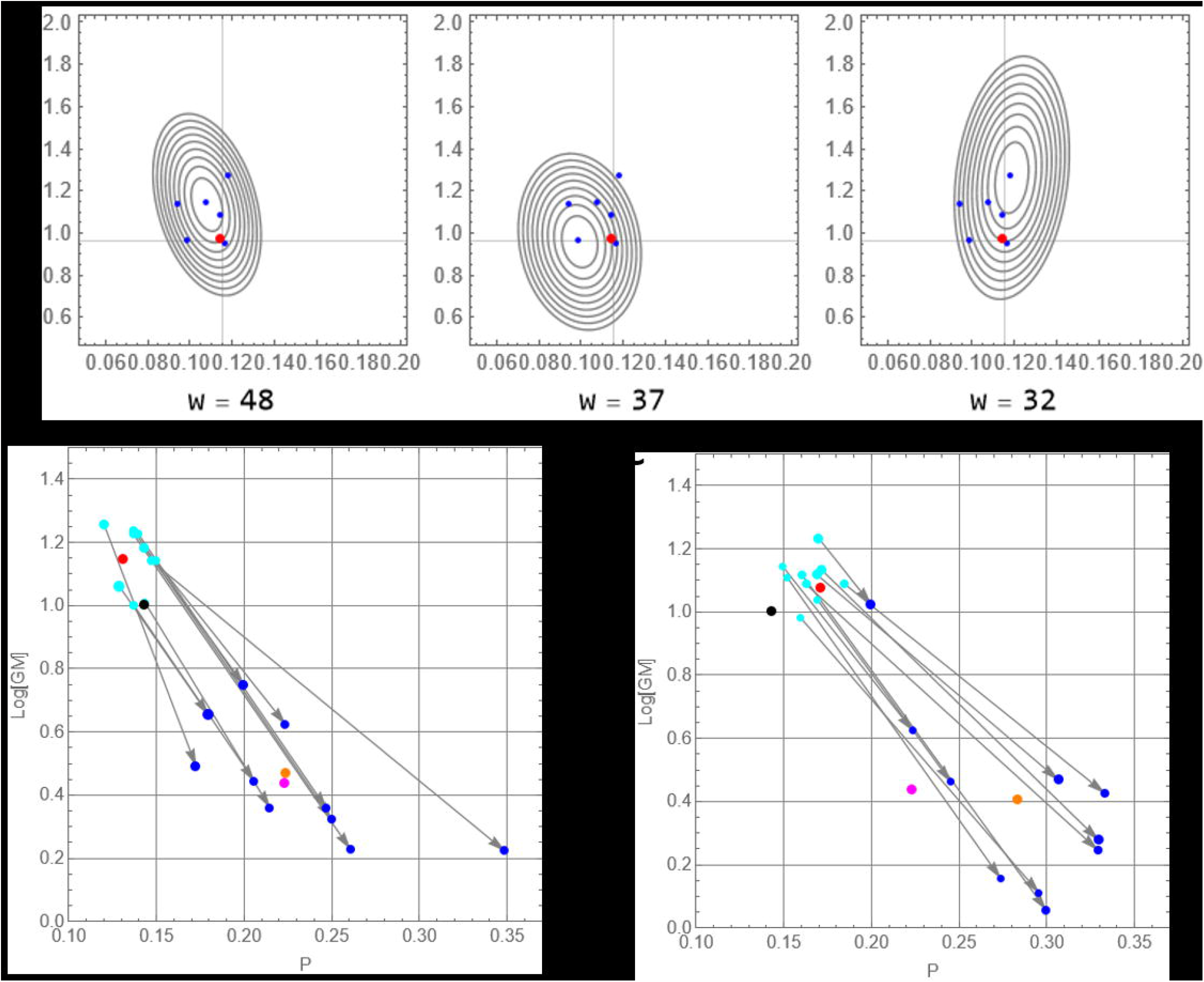
(a) Local SCORE cluster {*z* _*j*_} (blue centers) of reference point *z*_0_ (red), and three selected clouds with likelihood weights{*w*_*j*_}. Panels (b-c) show cluster-based reconstruction of PG-truth for a red reference *z*_0_, drawn from a SCORE community test (black cloud center): local cluster (cyan), is shifted to SCORE ‘truth’ (blue), and its weighted mean (orange) serves to estimate true PG for red reference. Estimated PG is compared to SCORE truth (magenta).

A local SCORE-cluster serves to generate a synthetic SCORE-like community for a given single-slide *T*, with different {*T*_*j*_ : *j* = 1,…, *m*} contributing in proportion to their likelihood weights, ∑ _*j*_ *w*_*j*_ = 1. Many relevant statistics can be extracted from such SCORE-like composites.

For instance, a composite PG-cloud of *z*_0_ could be generated from weighted SCORE ensembles, and its center *z*_*C*_ estimated via weighted sum of the constituent center, *z*_*C*_ = ∑_*i*_ *z*_*i*_*w*_*i*_. Other statistics of *T* could be inferred from its SCORE-composite, including graded prevalence, as well as ‘truth reconstruction’.

### H. True prevalence-intensity reconstruction

Given a single-slide community test *T* with reference point *z*_0_ = (*P*_0_,*G*_0_), we select its local SCORE cluster, and link each constituent cluster center *z*_*i*_ to its SCORE-truth, *Z*_*i*_ weighted via rescaled likelihood value, *w*_*i*_ ∝ *D*_*i*_ (*z*_0_), ∑_*i*_ *w*_*i*_ = 1. The proposed cluster-estimate of true PG is given by weighted mean,

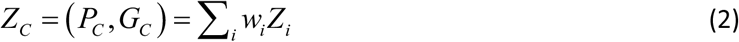

We illustrate the procedure for specific test data in Figure 3 (b-c). Two reference points

*z*_0_ (red) are extracted from resamples of two SCORE communities. In both cases, local SCORE-cluster centers (cyan) are shifted towards their true PG-values (blue), whose weighted mean (orange) gives cluster truth, *Z*_*C*_. As both reference points *z*_0_ come from SCORE resamples (could centers marked in black), we can compare ‘cluster truth’ (orange) with the ‘SCORE truth’ (magenta). In Figure 3 (b), two estimates come fairly close, so SCORE-cluster gives a good approaximation of the ‘truth’. In panel (c) two estimates are further apart (so cluster*Z*_*C*_ oveseatimates *Z*_*T*_), which is partly due to position of selected reference (red) relative to the SCORE center (black). We also note that ‘red resample’ (b) has higher likelihod than (c).

### I. Model validation

To validate our model, we applied it to SCORE community data, and an additional collection of non-SCORE communities with mixed test data. In each case, a single-slide random snapshot (*T*) was drawn from mixed data, and the clustering methodology applied to the reference values *z*_0_. Cluster outcomes, particularly estimated ‘true’ *P*_*C*_ was compared to the source ‘truth’ *P*_*T*_. In both cases, SCORE and non-SCORE, the truth is available via mixed (multi-day) test averaging.

In general, we should not expect a perfect match between *P*_*T*_ and *P*_*C*_, as shown in the previous section. So, our validation scheme aims to assess statistical robustness of cluster methodology across the entire community span.

We proceed by selecting a reference point *z*_0_ for each sampled community, and generating its local SCORE-cluster{*z*_*i*_ : *j* = 1,…, *m*}, along with the ‘source truth’{*Z* _*j*_}, and likelihood weights {*w*_*j*_}. The resulting cluster *Z*_*C*_ = (*P*_*C*_, *G*_*C*_), equation (2), is compared to the ‘source truth’ *Z*_*T*_ = (*P*_*T*_, *G*_*T*_). The scatter plot (*P*_*T*_, *P*_*C*_) is shown in Figure 4 (b); it includes 1624 SCORE testbed (gray points), and 7 non-SCORE communities (blue). Figure 4 (b) can be contrasted to ‘raw-data resampling’ of Figure 4 (a). The latter (raw resampling) exhibits systematic downward bias (gross underestimation of truth) and high variability. The former (cluster selection) is closer to diagonal ‘truth’ with narrower dispersal. Figure 4 (c) illustrates this gap by cross-sectional (vertical) slices of two distributions (a-b) at a fixed value *P*_*T*_ = 0.5 .

**Figure 4:**
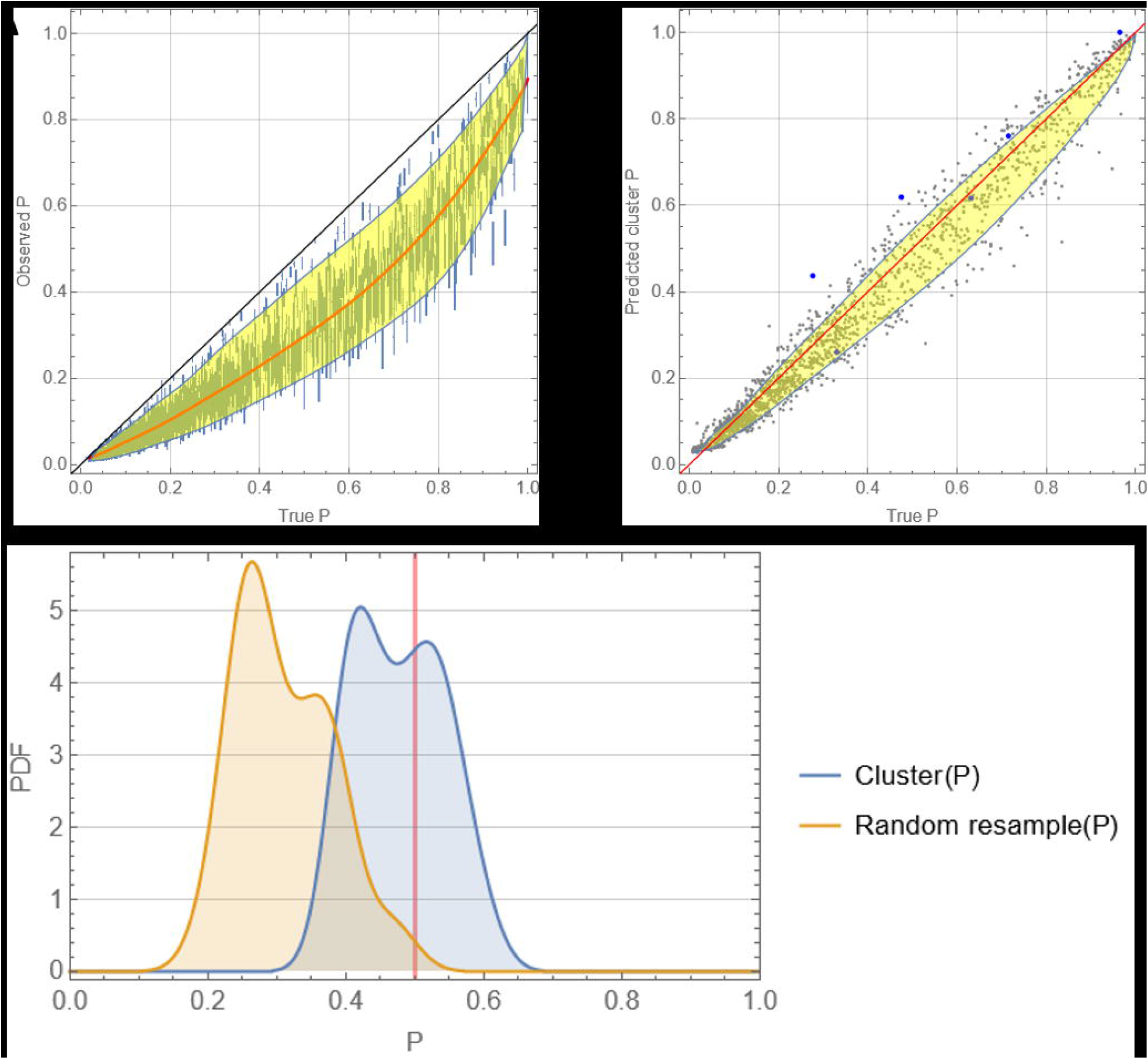
Panel (a) raw-resample errors bars of Figure 2(b) augmented with quantile marginal curves as functions of true *P*_*T*_. Panel (b) scatter plot of cluster selection scheme (*P*_*T*_, *P*_*C*_) for combined dataset made of 1624 SCORE values (gray dots) and additional communities (blue dots), augmented with quantile marginal curves. Panel (c) shows cross-sectional snapshot distributions of panels (a)-(b) at a fixed *P*_*T*_ = .5 : raw resample (yellow) vs. cluster (blue)

In both cases (panels (a)–(b) of Figure 4), we approximated discrete data-point (mean + uncertainty) by smooth functions (blue curves) that mark 5-95% quantile ranges of vertical scatters, in different P_T_ -prevalence bands. Those functions could serve as crude markers of predicted ‘true ranges’ for single-slide community snapshot. But wide marginal ranges, particularly panel (a), have little predictive value for prevalence assessment. To illustrate this point, we take a single-slide (observed) prevalence *P* = 0.4 ; panel (a) would predict ‘true range’ (0.48 < *P*_*T*_ < 0.78) –well above ‘observation’ with wide margins of uncertainty, while panel (b) would narrow the range (0.35 < *P*_*T*_ < 0.5) – closer to predicted cluster ‘truth’.

Overall, blue margin-curves of (b-c) rely on prevalence estimates alone, and give widely uncertain predictions. The key advantage of cluster-selection methodology is greater accuracy of ‘true’ prediction and reduced uncertainty, achieved via combined P-G statistics.

### J. P-G estimates of truth

An ideal tool for program managers would be a simple ‘function’ or ‘numeric code’ that would take a raw-data input, e.g. observed (*P,G*) -values, and predict ‘true’ prevalence *P*_*T*_ = *f* (*P,G*), within error margins. A version of such function (‘pocket chart’) was proposed by de Vlas [5] using a statistical model of test EPG, made of negative-binomial (NB) distributions. It assumed a hypothetical worm-burden stratification of host communities, and egg-release process by host-strata, described by suitable NBs. True *P*_*T*_ in this model, corresponds to ‘positive worm-burden’, while ‘egg-prevalence’ (test observation), *P* < *P*_*T*_, is comprised of ‘positive egg-release’ fractions of infected strata. He computed model function *f* (*P,G*) ; de Val et al. [5] shows its iso-contours in the (P,G)-plane, for a range of P_T_ –values (de Vlas topography). Our SCORE-cluster methodology yields an empirical version of ‘pocket chart’ (Figure 5). It resembles qualitatively de Vlas ‘pocket chart’, but SCORE ‘topography’ built on discrete cluster selection, makes it more rugged compared to [5]. However, our methodology goes beyond topographic chart of Figure 5. We developed a simple and efficient computer tool (SCORE calculator), that would any take any raw test-data, and estimate its ‘true’ statistics within uncertainty margins.

**Figure 5:**
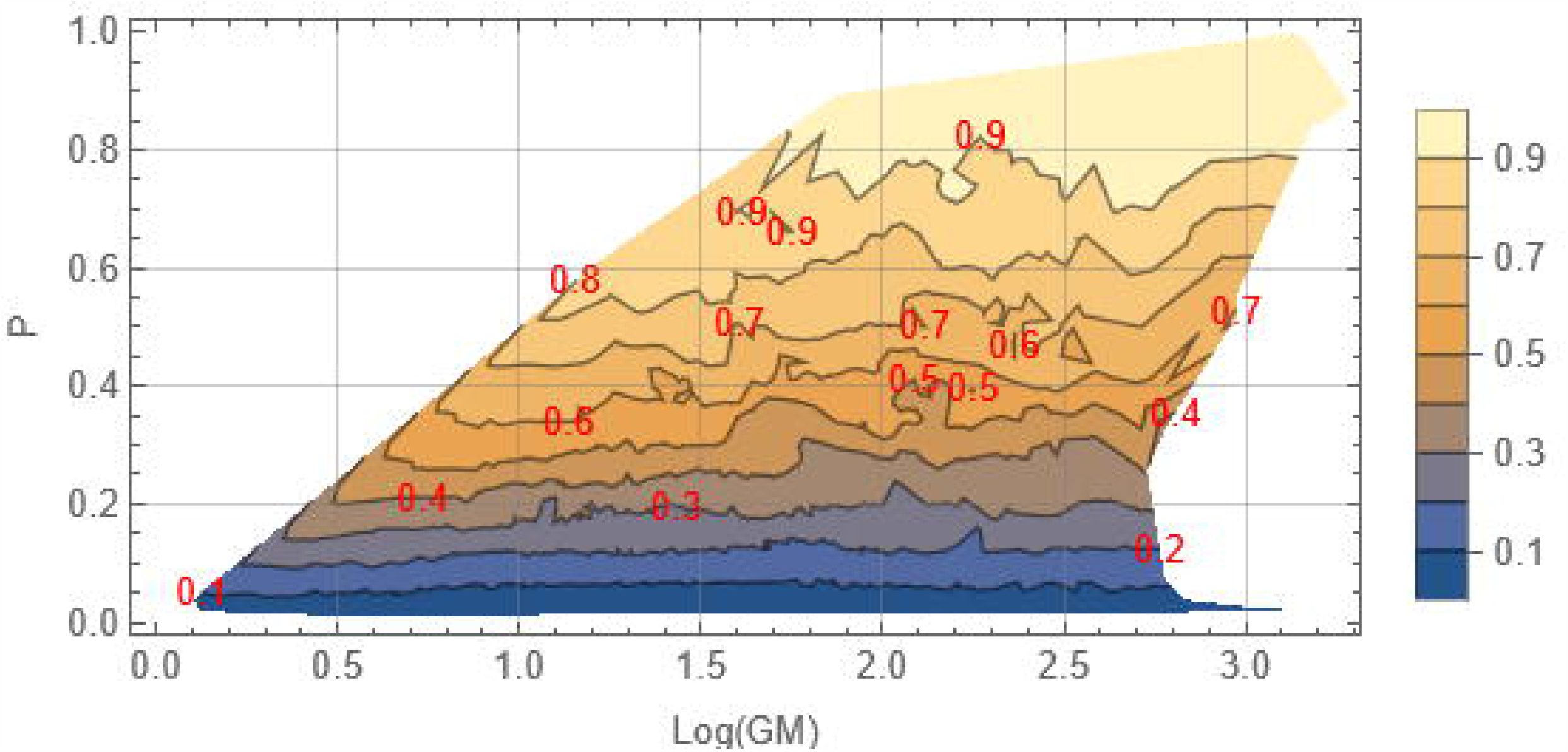
SCORE version of de Vlas topography in the G-P plane (pocket chart of [5]); red iso-contours mark fixed values of true *P*_*T*_ = *f* (*G, P*), as function of (G,P)

### K. Beyond prevalence-intensity

Prevalence serves as a key measure of community infection widely used in control programs, see e.g. WHO roadmap strategies [18]. Another important statistic proposed by the WHO is the graded prevalence based on EPG-counts: 0 < EPG < 100 (Light), 100 < EPG < 400 (Moderate), 400 < EPG (Heavy). We call them{*L, M, H*}; the latter (*H*) serving as a proxy of Schistosomiasis morbidity (heavy infections are often correlated with chronic conditions). Thus, WHO control strategies rely not only on the combined prevalence, *P* = *L* + *M* + *H*, but include *H*, as specific target.

The SCORE-cluster methodology allows one to assess different community statistics, including LMH. As above, a single-slide community test would give specific LMH-values, but a multi-slide (mixed) test would generate an LMH ensemble via resampling, alongside ‘true LMH’, based on average EPG scores (1). As above, one should not expect resampled LMH match the ‘truth’. Figure 6 (a) illustrates LMH discrepancies for a specific community test. Here resampled *L*; *M* underestimate ‘truth’, while *H* exceeds it.

**Figure 6:**
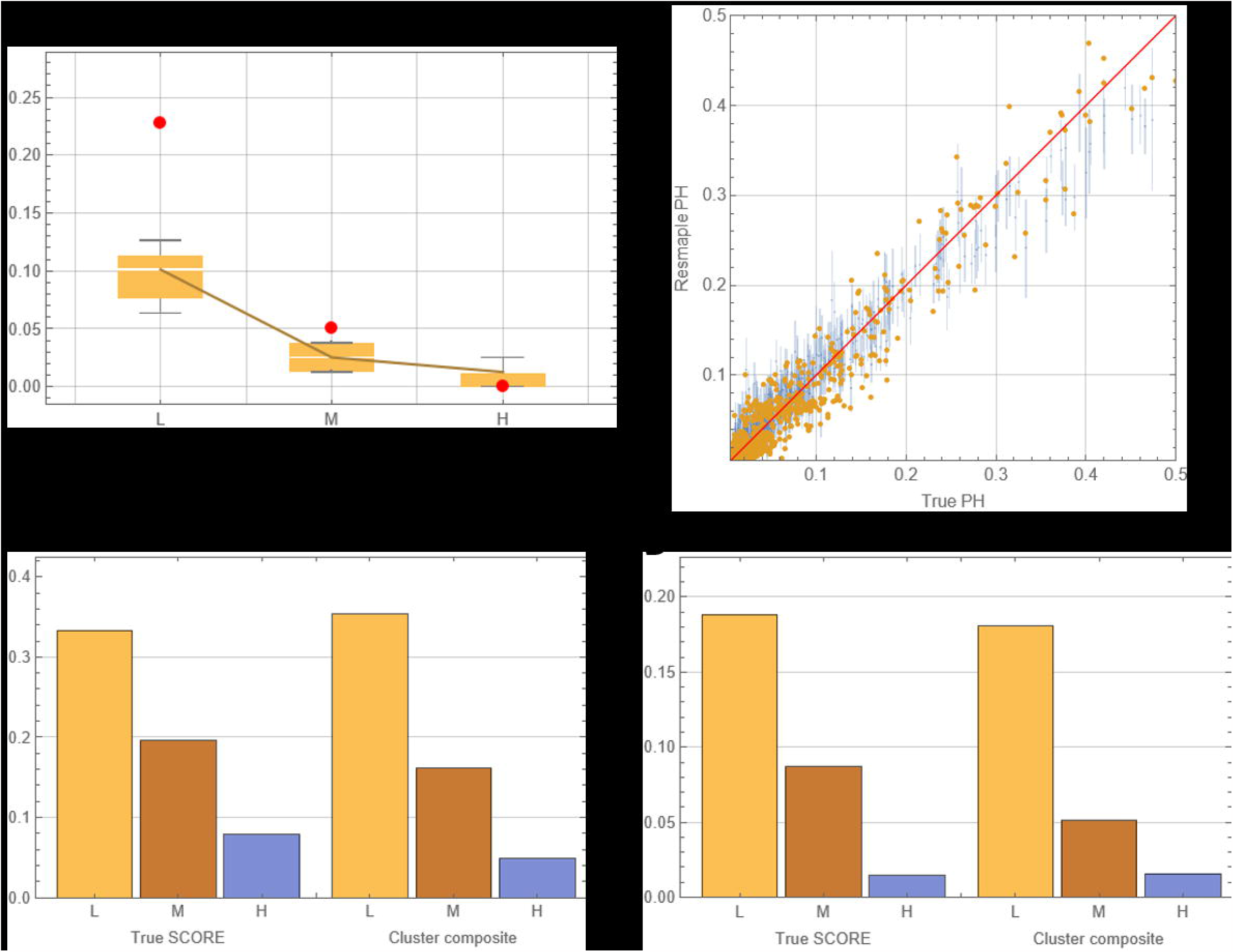
Estimates of graded Light-Moderate-Heavy (LMH) prevalence. Panel (a) compares LMH ensemble distribution (box-whiskers) against true LMH (red dots) for a specific community test. Panel (b) shows heavy prevalence *H* for 1620 community tests: bars correspond to raw-resample; yellow dots are cluster-composite estimates of *H*. Both are plotted against true SCORE *H*. Panels (c-d) LMH prevalence values (true vs. cluster composite) for two selected community tests.

The cluster-selection for LMH proceeds as above; starting with a PG reference *z*_0_ (raw test data), we generate its local SCORE-cluster{*z*_*i*_, *w*_*i*_ : *i* = 1,…, *m*}, along with their triplet ‘truth’ – a collection of mixed tests{*T*_1_;…;*T*_*m*_}. Each *T*_*i*_ has its true LMH(*L*_*i*_, *M*_*i*_, *H*_*i*_), weighted by PG-likelihood value *w*_*i*_ of *z*_*i*_. The resulting cluster estimate of LMH is given by the weighted sum

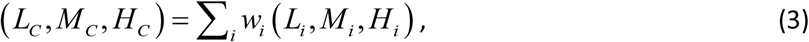

similar to equation (2). Indeed, each cluster test *T*_*i*_ contributes *w*_*i*_ -share of composite SCORE ensemble.

To assess the validity of cluster estimate (3) across all SCORE communities, we compared Predicted *LMH*_*C*_ with SCORE ‘truth’ *LMH*_*T*_, in particular heavy prevalence *H*. The scatter plot of Figure 6 (b) shows points (*H*_*T*_, *H*_*C*_) -yellow dots, with grey bars marking raw-resample estimates of *H*. As above, we do not expect a perfect match, but overall cluster selection exhibits higher accuracy and lower variability (yellow dots vs gray shaded area).

## Discussion

Conventional approaches to schistosomiasis surveillance and control rely on single-slide KK diagnostics. KK diagnostics exhibits high variability on individual (day-to-day) basis, and the resulting community assessment can be grossly underestimated relative to a putative multi-slide truth. To fill in the missing ‘observation-truth’ gap we propose to utilize an extensive dataset of multi-slide community tests of the SCORE project.

Our analysis combined conventional measures of community infection for mixed (multi-slide) tests (see e.g. [6, 7]-[8-10]), with alternative statistics derived from data resampling. We use consistently prevalence-intensity statistics; the latter measured by geometric mean (G) of positive counts. While prevalence P was the key target, we found P alone was insufficient for accurate assessment, but combined (P-G) provided essential tools for analysis; indeed, the entire scheme was set up and carried out in P-G (prevalence-intensity) plane (c.f. [5]).

The key application of our methodology is leveraging the SCORE dataset for any single-slide community test data T (SCORE or non-SCORE) to estimate its (unknown) true statistics, prevalence, intensity et al. This is accomplished by identifying a cluster of SCORE communities ‘similar’ to T, measured by the likelihood of generating T-statistics, via resampling. Once SCORE-cluster is identified, one can create a ‘virtual SCORE replica’ of community snapshot T, and infer its statistics, including unknown true (P-G), along with their uncertainty (error) margins.

The effect of combined (PG) statistics and cluster selection model are illustrated in Figure 4. Panel (a) derived from raw-data resampling (expected outcomes of single-slide KK screening) shows broad and consistently biased error-margins (well below true-P). Panel (b) derived by the SCORE-cluster scheme comes much closer to the truth with reduced error margins.

Our work highlights the role of combined P-G statistics for accurate community assessment, which could be relevant to other helminth infections. It also raises a challenging problem of developing P-G based control guidelines that would extend the current WHO-strategies based on prevalence alone [18]. Indeed, worm burden and the resulting egg-release, could vary widely within and between host communities. Intensity variable G complements prevalence P, as shown by SCORE data analysis (Figure 2). So, communities with near identical P could exhibit vastly different G-values (higher burden). They differ by EPG-count distributions or graded prevalence levels, like WHO LMH (Light-Moderate-Heavy) prevalence. Supplement Figure S3 demonstrates the effect of increased intensity G within a narrow prevalence band (*P* ≈ 0.2). Higher intensity (and associated worm burden) could affect other features in such, for instance their potential response to MDA control (see e.g. [8]). So control strategies that rely on prevalence alone could expect a mutitude of different outcomes.

Our work suggests that schistosomiasis control guidelines should account for P-G statistics to provide a more robust framework for disease control and elimination. However, the problem of extending WHO control guidelines would require not only a more detailed analysis of SCORE-like large-scale dataset (MDA response patterns), but also extensive numeric exploration of dynamic transmision models for SCORE-like communities (see e.g. [8, 9]).

### Limitations and extensions

The current version of cluster-selection scheme employs 1624 SCORE community tests, augmented with an additional 5-country dataset. It can be applied to any single-slide raw test data (reference point *z*_0_ = (*P*_0_,*G*_0_)) that falls within or near the lamina-shaped region of Figure 2 (a). Reference points outside this range may not produce statistically significant clusters for estimating ‘truth’. We do not know whether SCORE dataset covers all possible transmission environments and infection patterns, but it looks sufficiently representative in terms of prevalence values.

The proposed cluster methodology can be extended beyond SCORE testbed. Indeed, any mixed-test community data can be added to augment SCORE pool, like the 5-country dataset. Such extensions could only improve prediction skill and reduce model uncertainty.

Going beyond diagnostic assessment, our approach can be combined with dynamic transmission models ([8-10, 19]) for reanalysis of SCORE MDA-progress patterns, persistent hotspots, and efficient control strategies.

## Data and computer resources

The basic data source for our modeling and analysis is publicly available SCORE dataset www.clinepidb.org; it contains an additional file called ‘SCORE Five Country CCA Evaluation Cross-sectional’. The computer codes and procedures were developed and run on Wolfram Mathematica platform. Based on those, we deployed an open source SCORE-calculator. The code can take any raw KK-test input (EPG counts) for single or multiple communities, in any data format, and will output their ‘true’ statistics (PG and LMH) within error margins. This easy to use tool requires minimal experience in Mathematica, and is available at GitHub link: https://github.com/mln27/SCORE-Calculator

## Data Availability

All data produced in the present work are contained in the manuscript

## Acknowledgment

We thank our SCORE colleagues for providing SCORE datasets, and sharing their analysis and insights. We got many useful feedbacks from N. Lo (UCSF), and V. Mallikarjun (CWRU) has contributed to computer development.

## Authors contributions

DG and MLNM designed the project; DG performed the analysis and computer implementation; DG and MMLN interpreted the results; DG, MLNM and AM drafted the manuscript; DG, MLNM and AM reviewed and approved the final version.

## Ethics committee approval

NA.

## Patient Consent Statement

This study does not include factors necessitating patient consent.

