## Supplementary Material for "Improved assessment of *Schistosoma* community infection through data resampling methodology"

### Supplementary Materials

#### A. Sensitivity of KK diagnostics based on aggregate SCORE dataset

Here we provide a brief summary of KK diagnostic sensitivity based on the analysis of 210,000 individual SCORE tests. To this end we selected all multiple tests (doubles and triples) with at least one positive count (pool sizes $n_2, n_3$ respectively), and split each one of them into different 'positive-zero' mixture types,

$$
\begin{aligned}
n_2 &= n_2(0+) + n_2(++) \\
n_3 &= n_3(00+) + n_3(0++) + n_3(+++)
\end{aligned}
$$

True positivity rate, TPR = 1 – FNR (false negative ratio) can be estimated for each multiple pool (doublet, triplet) separately, or for the combined doublet + triplet pool. The results are shown in Table S1. It gives estimates of KK sensitivity (probability of detecting positive case from a single KK slide draw), for different multi-test data. The last raw of Table S1, does a similar exercise for double KK-draws from triplet tests. As expected, the latter has higher sensitivity

Table S1: KK sensitivity estimates

| | Type | TPR | TPR-value |
|---|---|---|---|
| Single slide draw | Doublet alone | $1 - \frac{n_2(0+)/2}{n_2}$ | 0.78 |
| | Triplet alone | $1 - \frac{n_3(0++)/3 + n_3(00+)2/3}{n_3}$ | 0.66 |

| | | | |
|---|---|---|---|
| | Combined | $1-\frac{n_2(0+)/2+n_3(0++)/3+n_3(00+)2/3}{n_2+n_3}$ | 0.67 |
| Double draw | Triplet | $1-\frac{n_3(00+)/3}{n_3}$ | 0.88 |

Reduced sensitivity of and high dispersal of KK-values of multiple tests are illustrated in Figure S1.

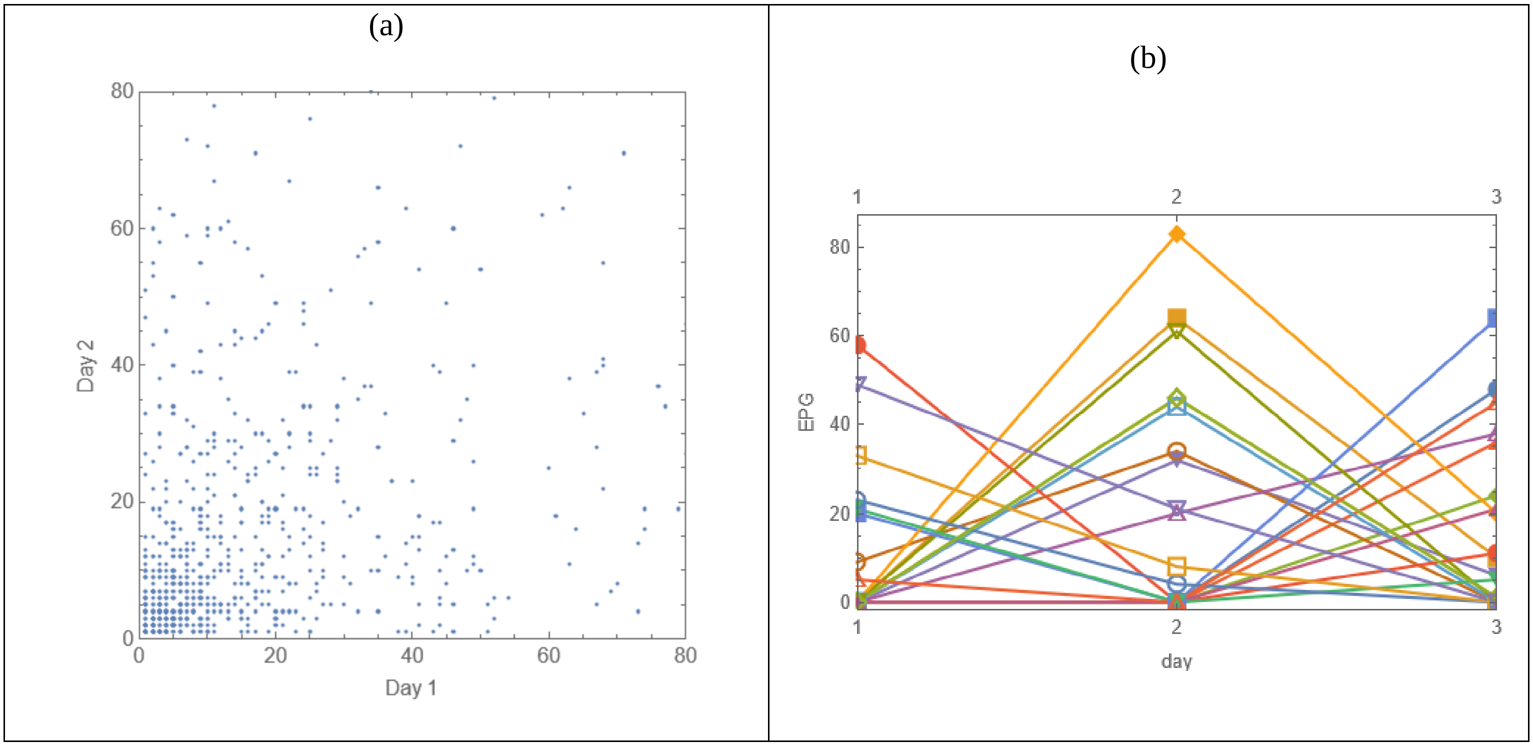

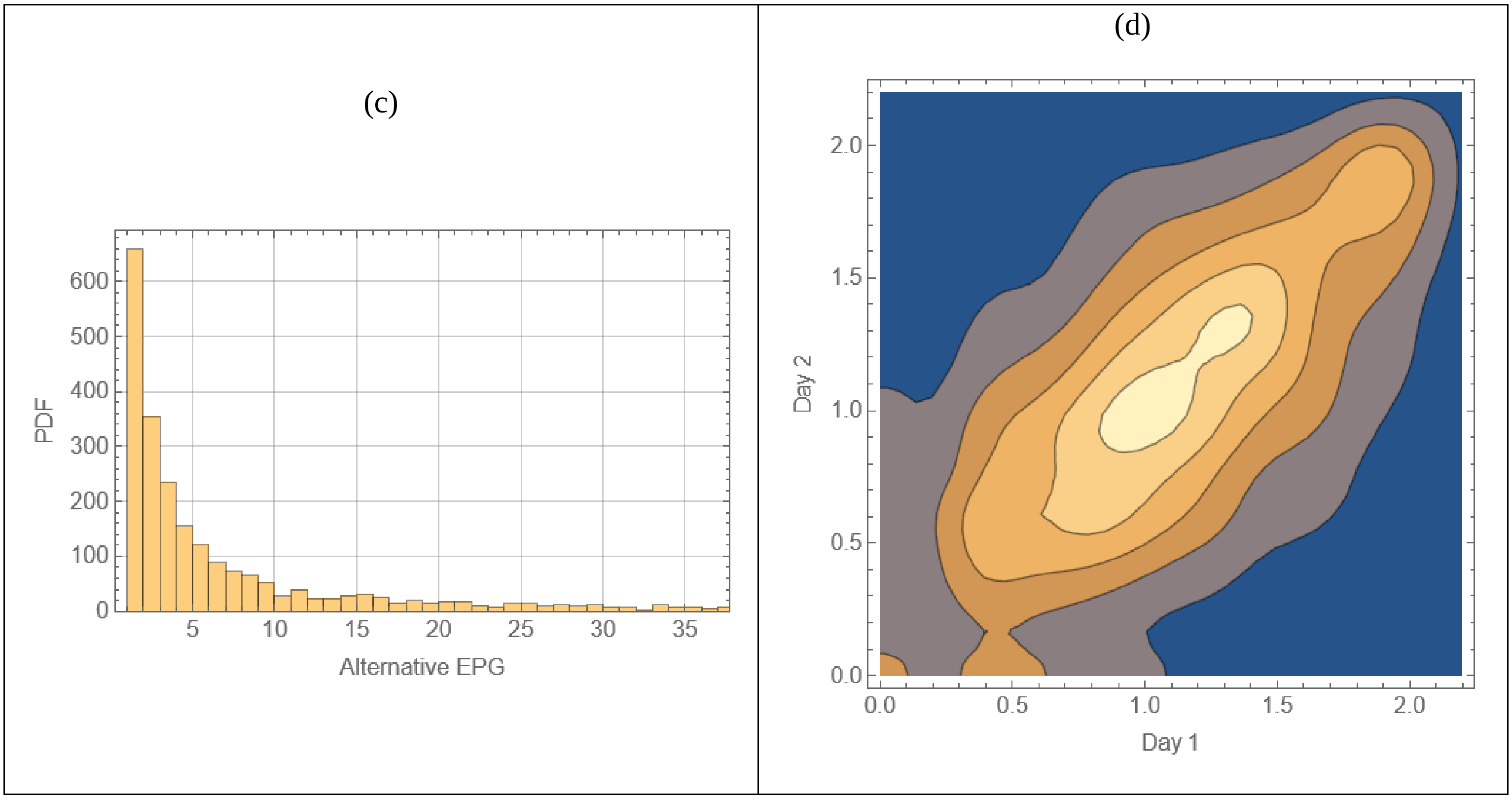


Figure S1: Multiple-stool test uncertainties: (a) random 800 double tests (day1-day2); (b) a random sample of triple tests with high day-to-day variability; (c) distribution of positive alternatives (e>0) for all (0,e) double types (over-dispersed pattern); (d) distribution density of all positive double tests $(e_1,e_2)>0$ on log-scale (the correlation coefficient = 0.59).

#### B. Mathematical and empirical background for P-GM methodology

P-GM methodology (prevalence + geometric mean) consist of replacing KK test data $(e_1,e_2,...)$, by a pair of statistics: P - fraction of positive counts ($e_i>0$), G – their geometric mean. Both measures extend naturally to weighted test-data $\{(e_i,w_i):i=1,...,n\}$. Here $w_0$ weighs e=0, $w_+=\sum_{e_i>0}w_i$, and P-G statistics are given by

$$P_w = \frac{w_+}{w};\quad G_w = \prod_{e_i>0} e_i^{\,w_i/w_+}$$

We consistently employ P–GM methodology on individual and communities levels. Specifically, for an individual (single-double-triple) test, its weight is equal to test multiplicity (1-2-3), and their P-G counts are shown in Table S2

Table S2: Weights and P-G statistics for single and multiple tests

| **EPG** | **Test weight** $w_T$ | **(G,P)** |
|---|---|---|
| $(0,0,a)$ | 3 | $(a,1/3)$ |
| $(0,a,b)$ | 3 | $(\sqrt{ab},2/3)$ |
| $(a,b,c)$ | 3 | $((abc)^{1/3},1)$ |
| $(0,a)$ | 2 | $(a,1/2)$ |
| $(a,b)$ | 2 | $(\sqrt{ab},1)$ |
| $(a)$ | 1 | $(a,1)$ |
| $(0);(0,0);(0,0,0)$ | m=1,2,3 | (0,0) |

The rationale for our ‘weighted’ choice is two-fold. On the one hand, it accounts for statistical significance of mixed EPG test (double, triple) vs. single test. On the other hand, splitting mixed types $(0+..)$ into ‘zero’ and ‘positive’ counts is consistent with raw-data resampling.

Here we provide a rational for using P-GM statistics for community assessment. We do it by examining some standard parametric distributions, e.g. negative binomial (NB), and by empirical community data drawn from SCORE.

A distribution $D(e)$ on nonnegative half-line ( $e \geq 0$ ), is split into its 'zero' and 'positive' components, $D(e) = p_0 \delta_0(e) + p_+ D_+(e)$, and $D_+(e)$ is transformed via log-change of variable $e \to \log(e)$. Such change converts geometric mean of $D_+$, into arithmetic mean of log-transformed $D_+$. Figure S2 illustrates it for several NB-distributions viewed as functions of continuous ( $0 < e < \infty$ ). In all cases, GM-value comes close to 'log-transformed' distribution peaks. Similar patterns could be derived mathematically for other types of half-line distributions.

For databased (empirical) distributions made of KK counts, log-change corresponds to log binning of test data. We demonstrate it in Figure S3 (b) for selected SCORE community tests within a narrow prevalence band about $P = 0.2$. Despite near identical P-values, the log-binned EPG-counts (empirical PDF), and the resulting graded-prevalence values (WHO Light-Moderate-Heavy) show distinct shift towards higher 'peak-location' (heavier infection burden) with increased G ('blue' vs. 'yellow' vs. 'green' clusters). As in Figure S2, estimated empirical GM come close to log-binned distribution peaks. In all cases, we observe a transition to with increased GM, in host-pools of near identical prevalence.

Thus, our P-G statistics could serve as a crude proxy of empirical PDF ('peak location' or 'severity') complementing the conventional prevalence P. Similar methodology was employed in [1-3].

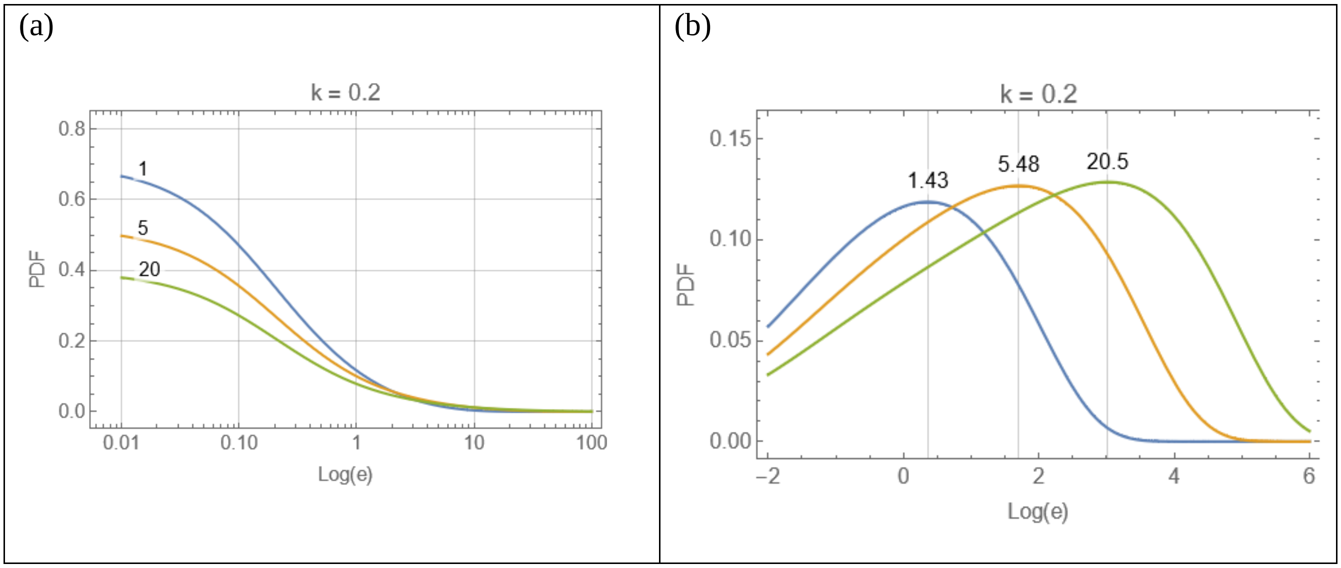


Figure S2: Continuous versions of $NB(E,k)$ for small k=0.2, and mean (expectation) values $E=(1,5,20)$. Panel (a) shows their PDF $P(e\,|\,E,k)$ in linear e (scaled logarithmically) – a featureless distribution pattern. Panel (b) shows the same PDF after log-variable change ( $\tilde{P}(u\,|\,E,k)=e^{u}P(e^{u}\,|\,E,k)$ , their u-peaks coinciding with GM-values of $\tilde{P}(\ldots)$-functions.

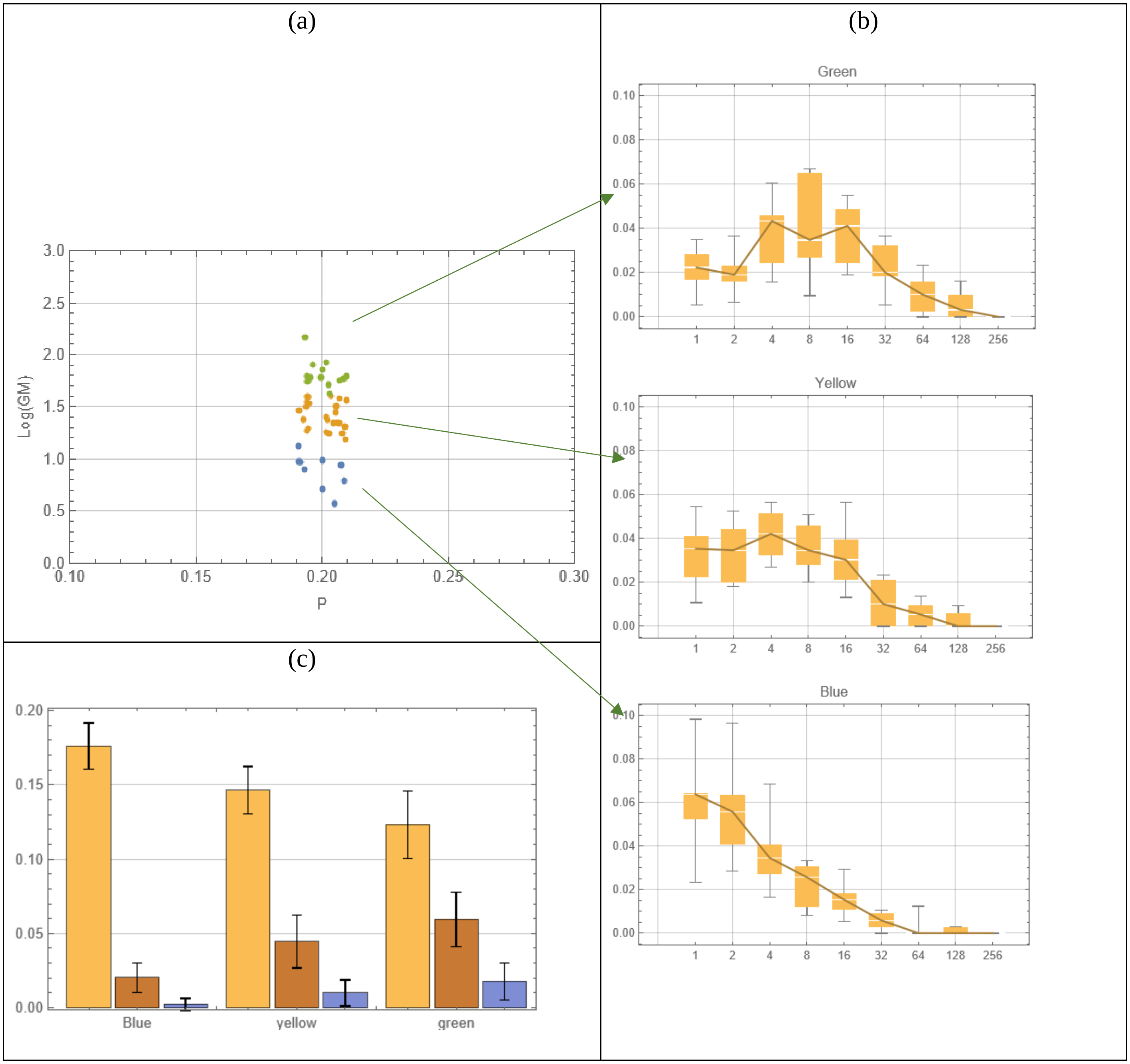


Figure S3: The effect of GM on EPG –distribution patterns. A narrow P-band of SCORE community tests range $|P-0.2|<.01$ is split into low (blue), middle (yellow) and high (green) GM-clusters. Their P-GM centers are shown in panel (a), and coarse (log-bin) c-PDF distributions (with quantile uncertainties) in panel (b). Panel (c) displays the WHO (Light-Moderate-Heavy) prevalence levels for 3 clusters (Lyellow, M- brown, H –blue). Panel (b) highlights a transition of EPG peak to higher values, panel (c) – consistent increase of heavy graded prevalence.


1. de Vlas SJ, Engels D, Rabello AL, Oostburg BF, Van Lieshout L, Polderman AM, Van Oortmarssen GJ, Habbema JD, Gryseels B: **Validation of a chart to estimate true *Schistosoma mansoni* prevalences from simple egg counts**. *Parasitology* 1997, **114 ( Pt 2)**:113-121.
2. Gryseels B, De Vlas SJ: **Worm burdens in schistosome infections**. *Parasitology today (Personal ed* 1996, **12**(3):115-119.
3. de Vlas SJ, Gryseels B: **Underestimation of *Schistosoma mansoni* prevalences**. *Parasitology today (Personal ed* 1992, **8**(8):274-277.
